# What Does the Public Want to Know About The COVID-19 Pandemic? A Systematic Analysis of Questions Asked in The Internet

**DOI:** 10.1101/2020.09.15.20192039

**Authors:** Jaiben George, Deepak Gautam, Vartika Kesarwani, PonAravindhan A Sugumar, Rajesh Malhotra

**Author notes:** **Correspondence:** Deepak Gautam, Department of Orthopedic Surgery, AIIMS, New Delhi, INDIA-110029.

## Abstract

**Background:** Quora is a popular question and answer (Q&A) website that enables people to connect with others and clear their doubts about the coronavirus disease (COVID-19). In this study, we analysed the content, type and quality of Q&As in Quora regarding this pandemic, and compared the information with that on World Health Organization (WHO) website.

**Methods:** We conducted a systematic search to include 964 questions in Quora. The tone of the question was categorized as either positive (questions with a primary intent to obtain information), negative (questions which represent panic or are related to misconception/false information) or ambivalent. The two most helpful answers of each question were graded for accuracy, authority, popularity, readability, and relevancy.

**Results:** 462 (48%) questions were classified as positive, while 391 (41%) were negative. Number of views were higher for negative questions (11421 vs 7300, p=0.004). Majority of the questions were on social impact (N=217, 23%), followed by politics (N=122, 13%) and disease management (N=96, 10%). Positive questions had more accurate, but less popular answers (p<0.05). Information related to 229 (28%) questions were present on WHO website, while partial information was present for 103 (11%) questions.

**Discussion:** Higher views with negative questions suggest that false and panic promoting information is more likely to get public attention. A substantial amount of questions was related to the present and future effects of COVID-19 on social and personal lives of the users which were not readily available on official health websites.

**Funding:** None

## Introduction

The novel coronavirus disease (COVID-19) originated in the Chinese province of Wuhan in December, 2019, and spread across the world over the next few months[1,2]. On March 11, 2019 WHO declared coronavirus as a global pandemic, at which point of time the epicentre of the disease had shifted from China to Europe[3]. Soon, countries across the globe were trying hard to control the pandemic with billions of people staying indoors as part of unprecedented measures to control the spread of the pandemic[4].

As COVID-19 spread rapidly, fear and the confusions surrounding it also spread[5,6]. With billions of people locked inside their home due to COVID-19, people are relying on Internet more than ever before to obtain information. Although the general public can obtain considerable amount of information about the disease from the websites of government agencies like World Health Organization (WHO), Centers for Disease Control and Prevention (CDC), Chinese CDC etc, these websites do not provide interactive opportunities for the public to seek answers to, or, answer specific questions. Such questions are often discussed in online question and answer (Q&A) platforms which have gained popularity in the recent times.

Quora is a popular Q&A website which enable users to raise their doubts which are then addressed by other internet users[7]. Q&A platforms allow users to ask versatile questions which may be of personal or public importance and are often answered by others who may or may not be expert of the content. Such websites have millions of followers, and continue to be an important platform for dissemination of information. For examples, the Coronavirus space in Quora alone has over 3.3 million followers as of May 13, 2020.The questions in Quora echo the voice of the public and understanding the questions asked by the users helps in identifying the concerns of the public from their perspective. Moreover, this understanding will help health authorities and governments to update their websites or other online platforms to cater to the needs of the public.

Therefore, we conducted a systematic analysis of Q&As in Quora. Specifically, we asked the following questions: 1) What are the type of questions asked in Quora? 2) What is the quality of answers to each of the questions? 3) How do the Q&As compare with the information in WHO official website?

## Methods

### Study Design

This was a cross sectional study of the questions asked in Quora about coronavirus from January, 2020 to March, 2020. These questions and answers were analysed for their content and quality in a systematic approach. Ethical approval was not required as this study was entirely based on data available in public domain and did not contain any protected health information.

### Search Criteria

Quora do not provide a specific search criterion to identify all the questions. As 100s of questions are being asked every hour with some of them being merged, we decided to analyse the most relevant 500 questions using each of the following search terms – ‘coronavirus’ and ‘COVID-19’. The top 500 most relevant questions from both the searches were compiled and duplicates removed to yield 965 unique questions. One of them was asked in 2018 and was excluded, resulting in 964 questions that were included for the final analysis in the study.

### Data Collection

The following information was collected for each question: date the question was asked, number of views, demographic details of the person who asked the question, and number of answers for the question. The country of the questioner was classified into one of the six WHO geographical regions. The tone of the question was analysed and each question was categorized as either having a positive (questions with a primary intent to obtain information) or negative tone (questions which represent panic or are related to misconception/false information). For example, a question about mortality of COVID-19 such as “How many people die of COVID-19?” was classified as positive while a question framed as “Are we all going to die from coronavirus?” was classified as negative. Questions which were not classified as positive or negative were considered to be ambivalent. The content of the question was also analysed and grouped under one of the following: virology (related to the nomenclature, origin, cause of COVID-19), symptoms, transmission, management (diagnosis, treatment, prevention), outcomes, myth (false information, misconceptions, conspiracy theories, etc.), social impact (impact on daily lives with respect to education, economy, lifestyle, etc.), politics (government policies, political parties or persons),statistics, and miscellaneous.

As all users are free to provide an answer to a given question, multiple answers are often available for each question. Quora ranks answers on a page according to how helpful they are after taking into account a variety of factors such as previous answers written by the author, experience of the author, whether the author is an expert in the subject, upvotes and downvotes (a measure of popularity of answers among the users) on the answer, etc[8]. We analysed the top 2 most helpful answers (or the top 2 ranked answers) for each question on a 3-point Likert scale using components of the DISCERN instrument in a modified manner[9]. The components were accuracy (based on the accuracy of the content of the answer as compared with WHO or CDC sources; 0-not accurate,1-somewhat accurate, 2-accurate), authority (based on the expertise or qualification of the person answering the question; 0-no expertise in the field,1-some expertise in the field, 2- an expert in the field),popularity (based on number of upvotes; 0-low [0-9 upvotes], 1- moderate[10-99 upvotes], 2- high[100 or more upvotes]), readability (whether a given answer is easy to read and comprehend for a lay person; 0- poor,1-fair, 2- good), and, relevancy (based on how relevant is the answer to the question; 0-not relevant,1-somewhat relevant, 2- relevant). The sum of the scores of each parameter (total score = 10 [5 × 2]) was used to assess the overall quality of the answer. All the Q&As were independently reviewed by two reviewers (VK, PS) for the tone, content, and quality, and any disagreement was resolved by a third reviewer (JG).

### Analysis

The Q&As in Quora were also compared with the information provided by WH0[10]. WHO has a dedicated FAQ section for COVID-19 on their online platforms as well as a separate section for answering the common myths related to COVID-19. The presence of information on WHO was also classified on a 3-point Likert scale as present, partially present or absent. Due to the positive skewness of the data, non-parametric Kruskal-Wallis test or Wilcoxon Rank Sum test was used. A post-hoc Dunn’s test was used to assess between the group differences. Categorical variables were analysed using a Chi-squared or Fisher Exact test. A p-value of less than 0.05 was taken as the threshold for statistical significance. All analyses were performed using R software (version 3.1.3, Vienna, Austria)[11].

## Results

The median number of views of the questions was 9202 (Range = 5.4 million to 164) with 16 (1.7%) questions having more than 1 million views. The ten most viewed questions are given in **Table 1**. Majority of the questions were asked by users from Region of the Americas (N=325, 34%), followed by South-east Asian region (N=199, 21%) and European region (N=115, 12%). Of those with gender information available (N=889, 92%), males had asked 700 (79%), while females asked 189 (21%). Overall, 462 (48%) questions were classified to have a positive tone, 391 (41%) were classified to have a negative. while 111 (12%) were found to be ambivalent. The characteristics of questions based on the tone are given in **Table 2**. The median number of views were higher in the questions with a negative tone (11421 vs 7300, p=0.004). Majority of the questions were on social impact (N=217, 23%), followed by politics (N=122, 13%) and disease management (N=96, 10%).

**Table 1.** The top 10 most viewed questions in Quora.

| No. | Questions |
| --- | --- |
| 1 | How has the coronavirus affected you? |
| 2 | How dangerous is the coronavirus? |
| 3 | When will the COVID-19 virus end? |
| 4 | How serious is the 2019–20 Coronavirus? |
| 5 | What does the coronavirus feel like? |
| 6 | Is COVID-19 likely to be at pandemic proportions for two years? |
| 7 | If the coronavirus has a 95% recovery rate, then what's with all the hype? |
| 8 | With the stock market down due to the COVID-19 coronavirus, is it a good time to buy or sell your stocks? |
| 9 | What are the positive impacts of the COVID-19 crisis? |
| 10 | What would you do if you found out you had COVID-19 (coronavirus)? |

**Table 2.** Characteristics of the included questions based on the tone

| **Variable** | **Positive**<br>**(N=462)** | **Negative**<br>**(N=391)** | **Ambivalent**<br>**(N=111)** | **P-value** |
| --- | --- | --- | --- | --- |
| Number of views, median<br>(interquartile range) | 7300<br>(3240-24537) | 11421<br>(3017-56985) | 11332<br>(4441-55916) | 0.011 |
| Gender of questioner, n (%) |  |  |  | 0.257 |
| Male | 336 (72.7%) | 289 (73.9%) | 75 (67.6%) |  |
| Female | 96 (20.8%) | 67 (17.1%) | 26 (23.4%) |  |
| Not available | 30 (6.5%) | 35 (9.0%) | 10 (9.0%) |  |
| Region of questioner, n (%) |  |  |  | 0.006 |
| Region of Americas (N=325) | 150 (32.5%) | 134 (34.3%) | 41 (36.9%) |  |
| South-east Asian (N=199) | 122 (26.4%) | 62 (15.9%) | 15 (13.5%) |  |
| European (N=115) | 55 (11.9%) | 49 (12.5%) | 11 (9.9%) |  |
| Western Pacific (N=41) | 20 (4.3%) | 19 (4.9%) | 2 (1.8%) |  |
| African (N=9) | 1 (0.2%) | 8 (2.0%) | 0 (0%) |  |
| Eastern Mediterranean (N=7) | 3 (0.6%) | 3 (0.8%) | 1 (0.9%) |  |
| Not available (N=268) | 111 (24%) | 116 (29.7%) | 41 (36.9%) |  |
| Content of question, n (%) |  |  |  | <0.001 |
| Social impact (N=217) | 78 (16.9%) | 84 (21.5%) | 55 (49.5%) |  |
| Politics (N=122) | 51 (11%) | 48 (12.3%) | 23 (20.7%) |  |
| Management (N=96) | 67 (14.5%) | 28 (7.2%) | 1 (0.9%) |  |
| Virology (N=88) | 67 (14.5%) | 19 (4.9%) | 2 (1.8%) |  |
| Transmission (N=81) | 49 (10.6%) | 31 (7.9%) | 1 (0.9%) |  |
| Myth (N=75) | 2 (0.4%) | 70 (17.9%) | 3 (2.7%) |  |
| Outcome (N=65) | 32 (6.9%) | 29 (7.4%) | 4 (3.6%) |  |
| Statistics (N=60) | 44 (9.5%) | 14 (3.6%) | 2 (1.8%) |  |
| Symptoms (N=33) | 28 (6.1%) | 3 (0.8%) | 2 (1.8%) |  |
| Miscellaneous (N=127) | 78 (16.9%) | 84 (21.5%) | 55 (49.5%) |  |
| Number of answers per question,<br>median (interquartile range) | 11 (6-26) | 15 (7-33) | 14 (8-32) | 0.002 |

Except for 9, all the other questions were answered. 61 (6%) questions had more than 100 answers, with the median number of answers being 12 (interquartile range, IQR: 7-30). The mean overall quality of the top ranked answer was 5.9±1.5, while it was 5.5±1.5 for the second ranked answer. Highest mean scores were seen for the relevancy and readability sections (**Table 3**). Positive questions were more likely to have an accurate answer than negative questions (Answer 1: 1.3±0.6 vs 1.2±0.6, p=0.003; Answer 2: 1.3±0.6 vs 1.1±0.5, p<0.001) although the positive questions had less popular answers (Answer 1: 0.4±0.7 vs 0.6±0.8, p<0.001; Answer 2: 0.3±0.6 vs 0.5±0.7, p<0.001) (**Table 3**).

**Table 3.**
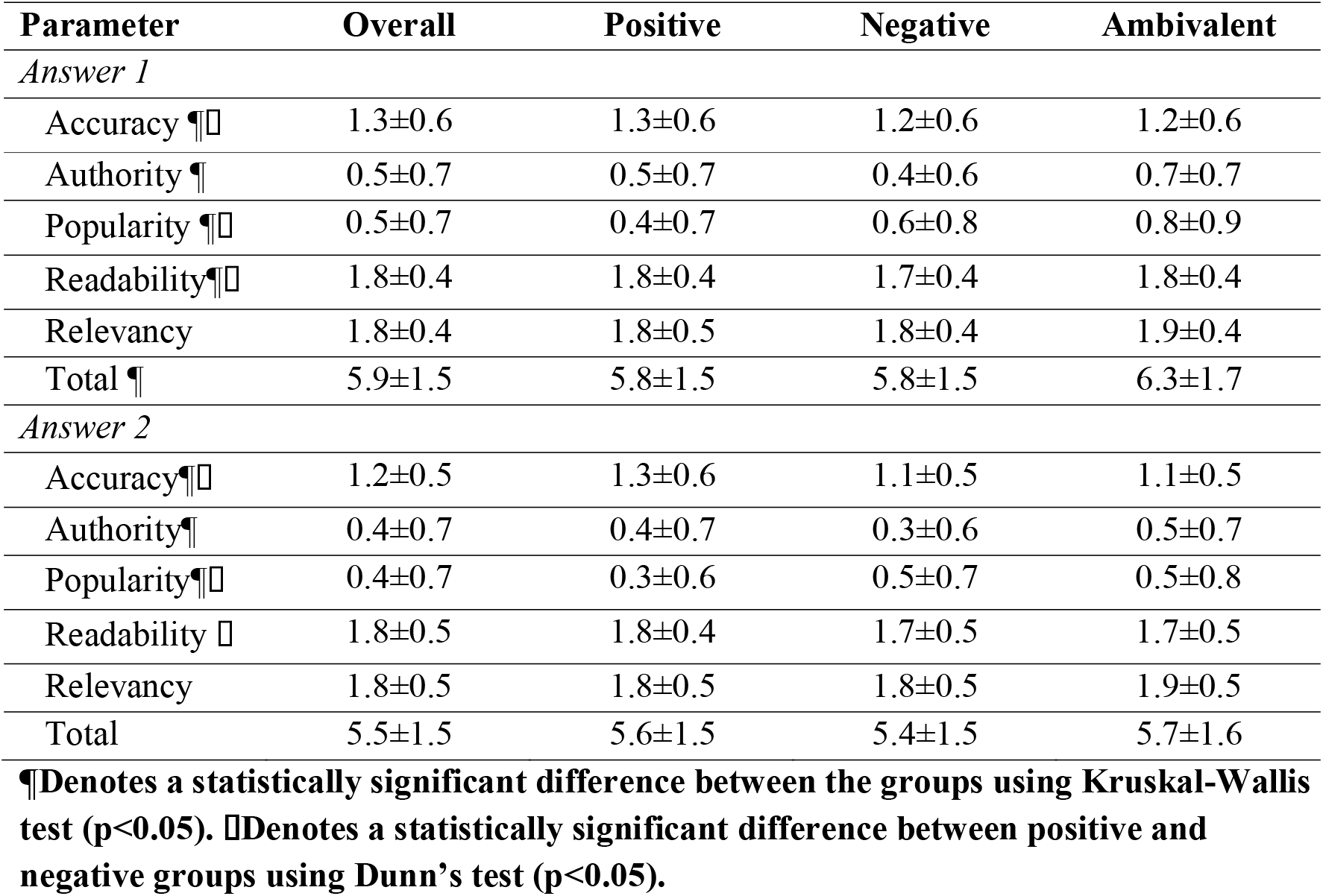
The quality of the answers based on the tone of the questions

| Parameter | Overall | Positive | Negative | Ambivalent |
| --- | --- | --- | --- | --- |
| *Answer 1* |  |  |  |  |
| Accuracy ¶□ | 1.3±0.6 | 1.3±0.6 | 1.2±0.6 | 1.2±0.6 |
| Authority ¶ | 0.5±0.7 | 0.5±0.7 | 0.4±0.6 | 0.7±0.7 |
| Popularity ¶□ | 0.5±0.7 | 0.4±0.7 | 0.6±0.8 | 0.8±0.9 |
| Readability ¶□ | 1.8±0.4 | 1.8±0.4 | 1.7±0.4 | 1.8±0.4 |
| Relevancy | 1.8±0.4 | 1.8±0.5 | 1.8±0.4 | 1.9±0.4 |
| Total ¶ | 5.9±1.5 | 5.8±1.5 | 5.8±1.5 | 6.3±1.7 |
| *Answer 2* |  |  |  |  |
| Accuracy ¶□ | 1.2±0.5 | 1.3±0.6 | 1.1±0.5 | 1.1±0.5 |
| Authority ¶ | 0.4±0.7 | 0.4±0.7 | 0.3±0.6 | 0.5±0.7 |
| Popularity ¶□ | 0.4±0.7 | 0.3±0.6 | 0.5±0.7 | 0.5±0.8 |
| Readability □ | 1.8±0.5 | 1.8±0.4 | 1.7±0.5 | 1.7±0.5 |
| Relevancy | 1.8±0.5 | 1.8±0.5 | 1.8±0.5 | 1.9±0.5 |
| Total | 5.5±1.5 | 5.6±1.5 | 5.4±1.5 | 5.7±1.6 |

¶Denotes a statistically significant difference between the groups using Kruskal-Wallis test ( $p<0.05$ ). □Denotes a statistically significant difference between positive and negative groups using Dunn's test ( $p<0.05$ ).

Information related to 229 (28%) questions were present in the WHO website, while partial information was present for 103 (11%) questions. The information regarding the remaining 632 (66%) questions was missing in the WHO website. The median number of views were similar for questions with or without information in WHO (Present= 8654 [IQR:3581- 25006], Partially present =7138 [IQR:2673-33215], Absent =10008 [IQR:3232-44478],p= 0.574). Positive questions were more likely to have complete or partial information in the WHO (Positive= 56%, Negative=17%, Ambivalent=5%; p<0.001) (**Figure 1)**. Majority of the questions related to symptoms (79%) and virology (76%) had either complete or partial information in the WHO website, while only a few questions related to social impact (11%) and politics (10%) had the required information in WHO website (**Figure 2**).

**Figure 1.**
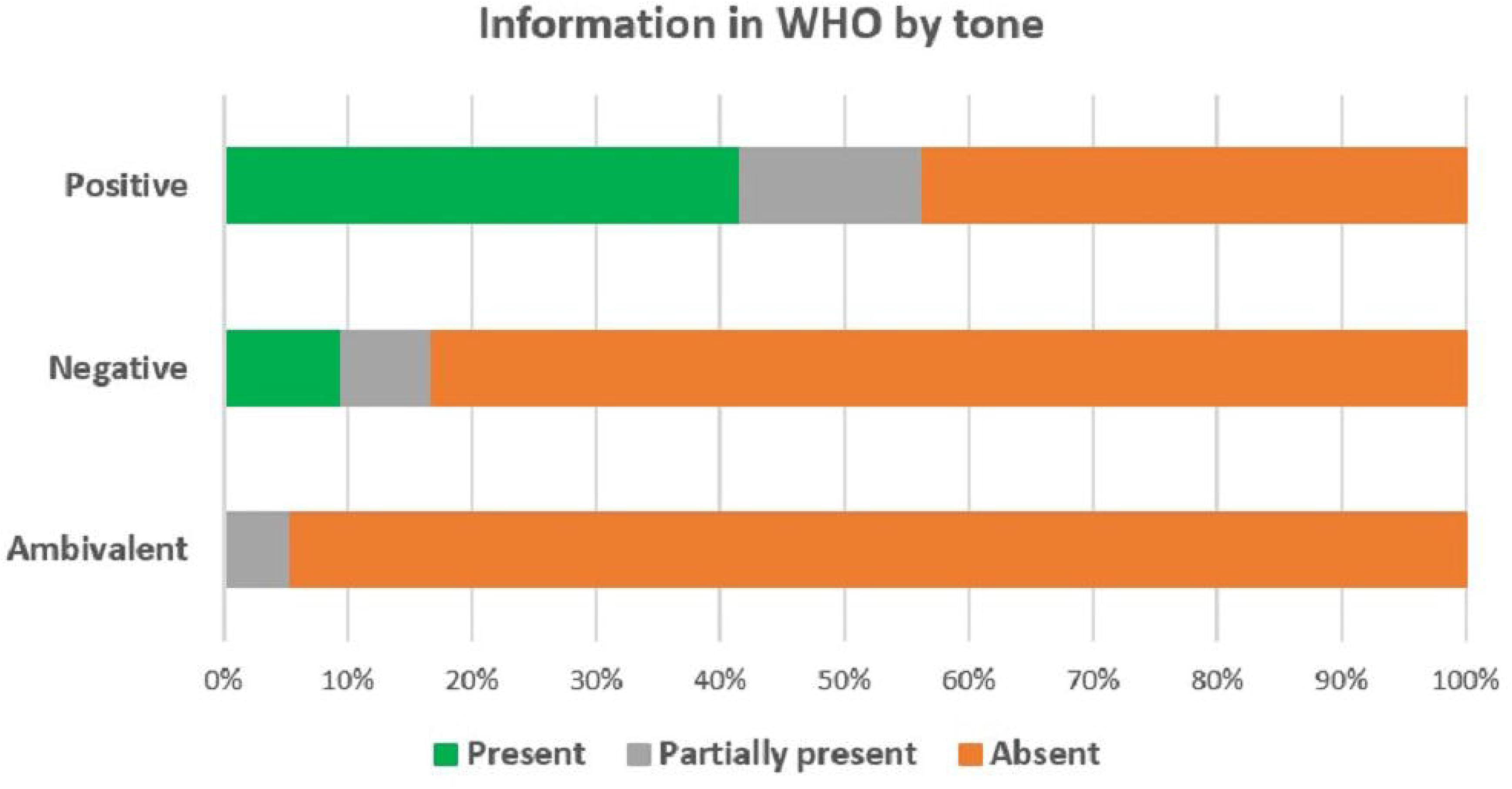
The presence of information in the WHO website based on the tone of the question.

**Figure 2.**
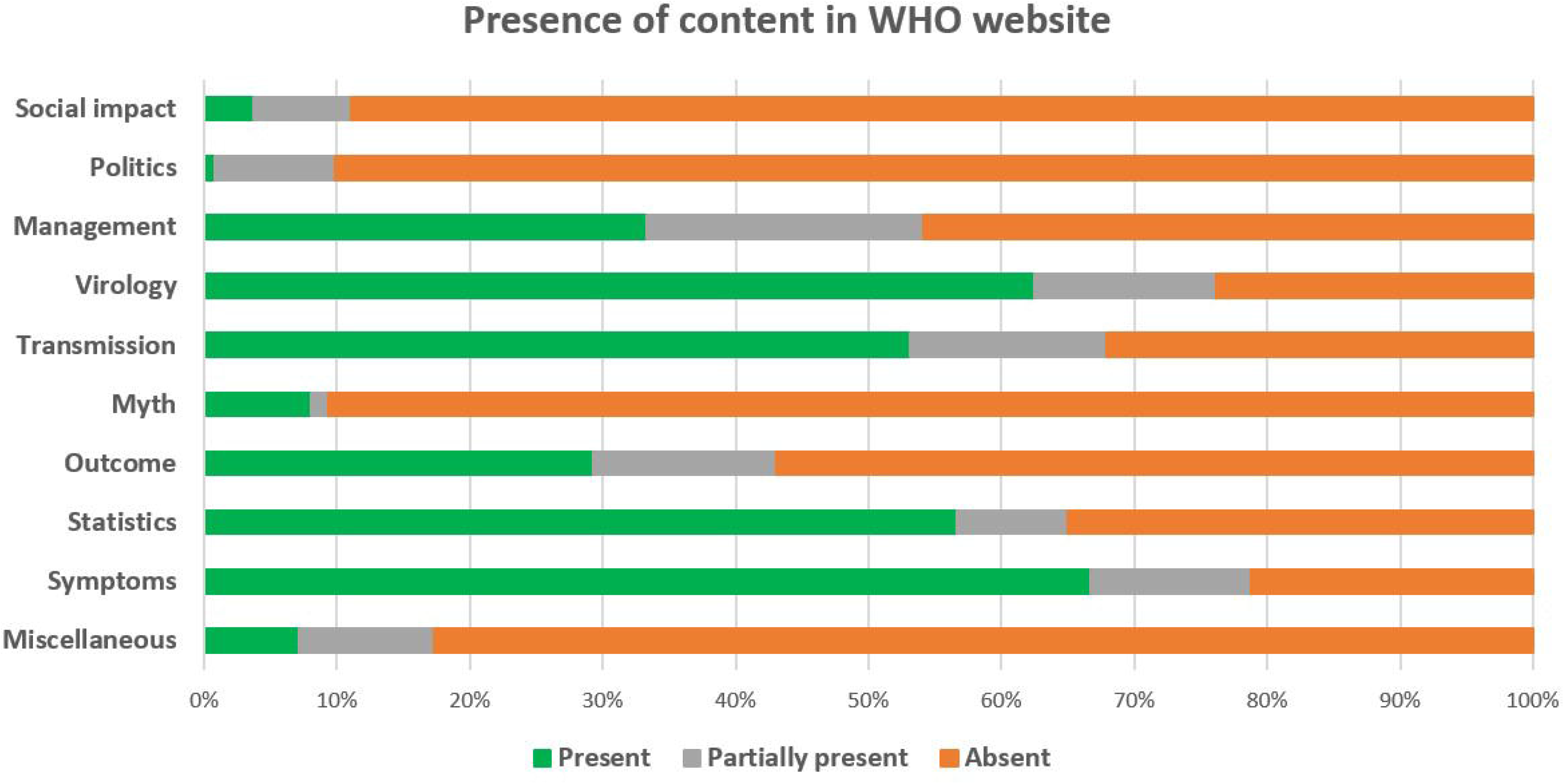
The presence of information in the WHO website based on the content of the question.

## Discussion

The COVID-19 pandemic has affected the lives of billions of people worldwide. Internet continues to be an important means to access information during this pandemic of unprecedented global impact. Quora provides people, many of them who are locked down in their homes, an opportunity to seek alleviation of their apprehensions about coronavirus. In this study, we analysed over 900 Q&As from Quora, and found that majority of them were related to the impact of coronavirus on their social lives. A considerable number of questions had a negative tone, and these questions had higher views, and had more popular and less accurate answers. WHO website was helpful in answering majority of the questions related to the medical aspects of virus. Nevertheless, a large number of questions related to the impact of COVID-19 on society were uncovered in WHO sources.

There are number of limitations to the study. Quora does not provide the option of a systematic search like various academic libraries such as PubMed or Embase. Moreover, the Q&As in Quora are updated at a rapid pace with 100s of questions added in every hour, and multiple questions being merged. As a result, a search conducted at a different time point might not have yielded the similar set of questions used in this study. However, this is unlikely to affect the results of the study as we used a large number of questions. Although three authors were involved in independently classifying the Q&As, the classification of questions as positive or negative might have been arbitrary. COVID-19 is an evolving situation, and as a result, the information related to COVID-19 in Quora and the information provided in WHO is also rapidly evolving, and might have affected the results of the study. The current study compared the information in Quora with that on WHO website only. Although WHO is an international body with its updates being followed globally, a number of other public health agencies and regional health bodies do provide reliable information about COVID-19 which may have been not present in WHO website. Finally, we only used the Q&As in English and the findings may be less applicable to non-English speaking nations.

Our study found that internet users from across the world ask a wide variety of questions in Quora with a number of questions having over a million views at the time of analysis. Majority of the questions were related to the social impact suggesting that people might be more concerned about the effect of COVID-19 on their personal and social lives than the medical aspects. A considerable number of questions had a negative tone. These questions had higher number of views suggesting that negative views on COVID-19 are likely to get more attention from the public. Prior studies have analysed the utility of different websites and online sources in providing information on various health topics. Keelan et al.[12] conducted an analysis of the content of YouTube about vaccination and found that about half of the videos posted were not supportive of immunization. Similar negative content was also observed in other studies evaluating health information in internet[13,14].

The present study reveals that Quora is a useful platform providing relevant and readable answers for most of the questions people have about COVID-19. Although most of the users were not experts in their respective fields, the most helpful answers appeared to be reasonably accurate. However, it was also found that answers to negative questions were less accurate and more popular suggesting that false information is more likely to spread in community which can lead to a state of panic. Such dissemination of negative information is often an unwanted consequence of the freedom of internet[13-16]. But, like most social medial platforms, Quora provides the users with the option of reporting content if it is inaccurate, not seeking a genuine answer, disparaging towards a person or a group, etc.

The information asked in about one-third of the questions was available in the WHO website. While information about the COVID-19 virology, transmission, management were more readily available in WHO, information about its impact on the society and government policies were not found in WHO. This may also explain the higher popularity of negative content in Quora as such topics may not be discussed freely in official health websites. Many questions in Quora reflected the frustration and helplessness in the society with the restrictions imposed as part of COVID-19 and a desire to see the end of it. Although the course of the pandemic is unpredictable, it might be helpful if health bodies and governments come up with possible estimates of the pandemic size and measures to relax restrictions at the earliest, and communicate this with the public. The inclusion of contents in WHO website such as ‘Coping with stress during COVID-19’ and ‘Staying active during COVID-19’ are commendable efforts in the direction of connecting with the public, and should be promoted by other agencies as well[17,18].

In summary, we found that a large number of users obtain information regarding COVID-19 from Quora. Majority of the questions had a positive tone although a considerable number of questions were of negative tone. The higher number of views with negative questions suggest that false and panic promoting information is more likely to be promoted through Internet. A substantial amount of questions in Quora were related to the present and future effect of COVID-19 on the social and personal lives of the users which was not readily available in online health information platforms like the WHO website. Considering the propensity of Internet to disseminate information (also misinformation) to a large and diverse audience, government and public health agencies should use Internet as an important tool to communicate with the public while monitoring its content.

## Data Availability

All the data used in this study has been obtained from Quora.

https://www.quora.com/

